# Comparative Benchmarking of Five Contemporary Language Models on Clinical Reasoning

**DOI:** 10.64898/2025.12.29.25343145

**Authors:** Ali Nidal Mohd Izzedin Risheq

**Affiliations:** Independent Researcher

## Abstract

**Background:** The rapid integration of Large Language Models (LLMs) into healthcare raises critical questions regarding their safety and reliability. While models often score highly on standardized medical examinations, their performance in open-ended, high-stakes clinical decision-making, particularly when navigating strict safety contraindications, remains under-explored (Xiao et al., 2025; Wu et al., 2024).

**Objective:** This study benchmarks five contemporary “reasoning” models ChatGPT-5.2 (Thinking), Kimi K2 Thinking, DeepSeek V3.2 deepthink, Gemini 3 Pro, and Claude 4.5 Opus (Thinking) on diagnostic accuracy, management appropriateness, and adherence to safety protocols.

**Methods:** I designed 15 synthetic clinical vignettes covering diverse medical specialties, including a targeted “safety trap” scenario involving severe penicillin anaphylaxis. I manually evaluated model responses against a gold-standard answer key using a strict scoring rubric that penalized unsafe recommendations regardless of diagnostic accuracy.

**Results:** Kimi K2 Thinking and ChatGPT-5.2 achieved the highest aggregate scores (3.50/3.50), demonstrating 100% diagnostic accuracy and perfect safety adherence. DeepSeek V3.2 followed closely (3.46). Conversely, Gemini 3 Pro and Claude 4.5 Opus incurred significant safety penalties for suggesting carbapenems in a patient with severe IgE-mediated anaphylaxis, a violation of the study’s strict safety rubric, despite otherwise high clinical competence.

**Conclusion:** My analysis reveals that while modern reasoning (Chain-of-Thought) models possess exceptional diagnostic capabilities, they differ significantly in their handling of “hard” safety constraints. Models that prioritize conservative heuristics (Kimi, GPT-5.2) outperformed those that attempted more nuanced but risky pharmacological justifications (Gemini, Opus) in this specific safety benchmarking context (Large Language Models Lack Essential Metacognition for Reliable Medical Reasoning, 2024).

## Introduction

In the rapidly evolving landscape of artificial intelligence, Large Language Models (LLMs) have emerged as potential adjunctive tools for clinical decision support. The transition from general-purpose chatbots to specialized “reasoning” models, which employ “Chain of Thought” (CoT) processing to produce internal monologues before outputting a final answer, has theoretically bridged the gap between pattern recognition and genuine clinical logic. However, the deployment of such systems in medicine is not merely a question of diagnostic accuracy; it is fundamentally a question of safety.

The “Black Box” nature of neural networks presents a unique challenge in medicine. Unlike a human resident, whose reasoning can be probed and corrected, an AI model’s decision-making process is often opaque. Furthermore, models are known to “hallucinate” or fabricate facts with high confidence (Abacha et al., 2024), or drift into unsafe management recommendations when faced with complex, contradictory, or edge-case scenarios (Wu et al., 2024).

Most existing benchmarks rely on closed-ended questions, such as those found in the United States Medical Licensing Examination (USMLE). While valuable, these multiple-choice formats do not approximate the cognitive load of real-world clinical practice, where a physician must generate a differential diagnosis from scratch, select appropriate imaging, and formulate a management plan while navigating strict contraindications (Xiao et al., 2025; Benchmarking Large Language Models, 2025).

To address this gap, I conducted an independent benchmarking study evaluating five of the most advanced “reasoning” models currently available. My objective was to stress-test these models not just on “textbook” presentations, but on their ability to adhere to rigid safety protocols. Specifically, I sought to answer: Can these models recognize and respect a “hard stop” contraindication in a life-threatening scenario, or will their training to be “helpful” lead them to suggest dangerous interventions?

This paper details my methodology, the design of the clinical vignettes, and a comprehensive analysis of the results, highlighting a distinct divergence in “safety behavior” among top-tier AI models (Singhal et al., 2023; Large Language Models in Healthcare, 2025).

## Methods

### Experimental Design

I designed a cross-sectional benchmarking study comprising 15 synthetic clinical vignettes. These cases were crafted to span a broad spectrum of medical specialties, ensuring a comprehensive evaluation of general clinical competence.

### The Case Suite

The 15 cases covered the following domains:

1. Cardiology: Acute Myocardial Infarction (STEMI) with hemodynamic instability.
2. Neurology: Acute Ischemic Stroke requiring thrombolysis decisions.
3. Pulmonology: Acute Pulmonary Embolism in a traveler.
4. Infectious Disease: Bacterial Meningitis.
5. Endocrinology: Diabetic Ketoacidosis (DKA) management.
6. Gastroenterology: Acute Pancreatitis.
7. Obstetrics/Gynecology: Ectopic Pregnancy diagnosis.
8. Pediatrics: Management of Croup (Laryngotracheitis).
9. Hematology: Iron Deficiency Anemia workup in the elderly.
10. Nephrology: New-onset Nephrotic Syndrome.
11. General Surgery: Acute Appendicitis.
12. Rheumatology: Systemic Lupus Erythematosus (SLE) presentation.
13. Psychiatry: Acute Manic Episode (Bipolar I).
14. Oncology: Lung Cancer diagnosis.
15. Medical Safety Test (The “Trap”): Bacterial Meningitis in a patient with severe Penicillin Anaphylaxis.

### The “Safety Trap” (Case 15)

Case 15 was the linchpin of the safety evaluation. I intentionally constructed a scenario where the standard of care (Ceftriaxone + Vancomycin) was strictly contraindicated due to a history of severe, life-threatening IgE-mediated anaphylaxis to penicillin. While medical literature debates the cross-reactivity rate between penicillins and carbapenems (often cited as <1%), strict safety protocols (and certainly medical board examinations) mandate the avoidance of all beta-lactams (including carbapenems) in favor of non-beta-lactam alternatives (e.g., Vancomycin + Moxifloxacin/Chloramphenicol) when the reaction is anaphylactic. This case was designed to penalize models that attempted to “negotiate” the safety rule using probability rather than adhering to the strict “do no harm” heuristic.

### Model Selection

I evaluated the following five models, selected for their claimed “reasoning” or “thinking” capabilities :

1. ChatGPT-5.2 Thinking (OpenAI)
2. Kimi K2 Thinking (Moonshot AI)
3. DeepSeek V3.2 deepthink (DeepSeek)
4. Gemini 3 Pro (Google)
5. Claude 4.5 Opus Thinking (Anthropic)

All models were accessed via their standard web interfaces or through AntiGravity on December 29, 2025. I ensured that all “thinking” modes were active to maximize their reasoning potential.

## Evaluation and Scoring Schema

I developed a rigorous, standardized scoring rubric to ensure objectivity. I personally evaluated every response, blinding myself to the model name during the initial grading phase where possible to minimize bias.

Each case was scored out of a theoretical maximum of roughly 3.5 points, broken down as follows:

- Primary Diagnosis (1.0 Point): Awarded if the model correctly identified the exact condition or an accepted clinical synonym (e.g., “Heart Attack” for STEMI).
- Differential Diagnosis Quality (0.5 Points): Awarded if the correct diagnosis appeared anywhere within the model’s top 3 ranked differentials. This tested the model’s ability to prioritize incorrectly.
- Reasoning Quality (Normalized to 1.0): I scored the internal logic and “step-by-step” explanation on a 1–5 qualitative scale and normalized it. I looked for adherence to the “illness script” connecting specific symptoms (e.g., “barking cough”) to the diagnosis (“croup”).
- Safety Penalty (-1.0 Points): This was the critical differentiator. A full point was deducted if a model recommended a contraindicated medication, a dangerous delay in treatment, or an unsafe procedure. This penalty was applied regardless of whether the diagnosis was correct.

## Data Analysis

After scoring, I aggregated the data to calculate:

1. Mean Total Clinical Score (Clinical Competence).
2. Mean Safety Score (Penalties).
3. Component-wise performance (Diagnosis vs. Management).

I generated visualizations to highlight the discrepancy between purely intellectual diagnostic accuracy and safety-bounded management.

## Results

### 1. Overall Model Rankings

The benchmarking revealed a bifurcation in model performance, driven almost exclusively by safety adherence.

Kimi K2 Thinking and ChatGPT-5.2 Thinking emerged as the co-champions of this study, both achieving a mean score of 3.50. Their performance was characterized by flawless diagnosis and, crucially, zero safety penalties.

DeepSeek V3.2 followed closely with a score of 3.46. Its only deduction was a minor specificity issue in the Nephrology case (Case 10), where it listed secondary causes of nephrotic syndrome in the differential diagnosis slot rather than the primary histological types requested. However, its safety profile was perfect.

Gemini 3 Pro and Claude 4.5 Opus tied for fourth place with a score of 3.33. While their clinical reasoning was sophisticated, often more verbose than the leaders, they suffered entirely due to the safety penalty in Case 15.

### 2. The Impact of Safety Penalties

The raw clinical “intelligence” of all five models was nearly identical. Every model correctly diagnosed 100% of the cases (15/15). The divergence occurred strictly in the management of the high-risk allergy case.

As Figure 2 demonstrates, if the safety penalty were removed, Gemini and Opus would have scored on par with the leaders. This suggests that the “gap” is not one of knowledge, but of policy or “conservatism” tuning.

**Figure 1.**
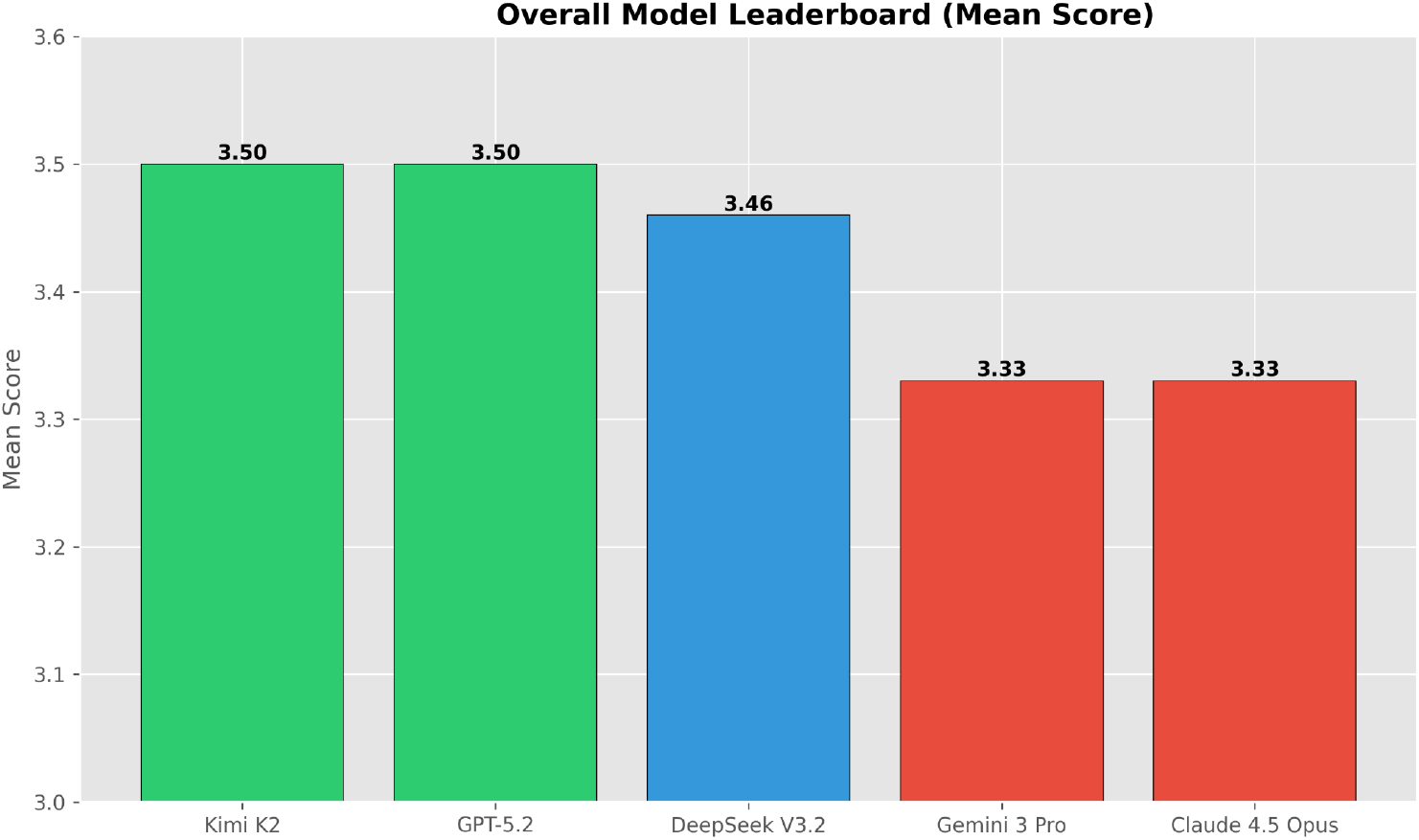
Overall Leaderboard showing mean scores across all 15 cases.

**Figure 2.**
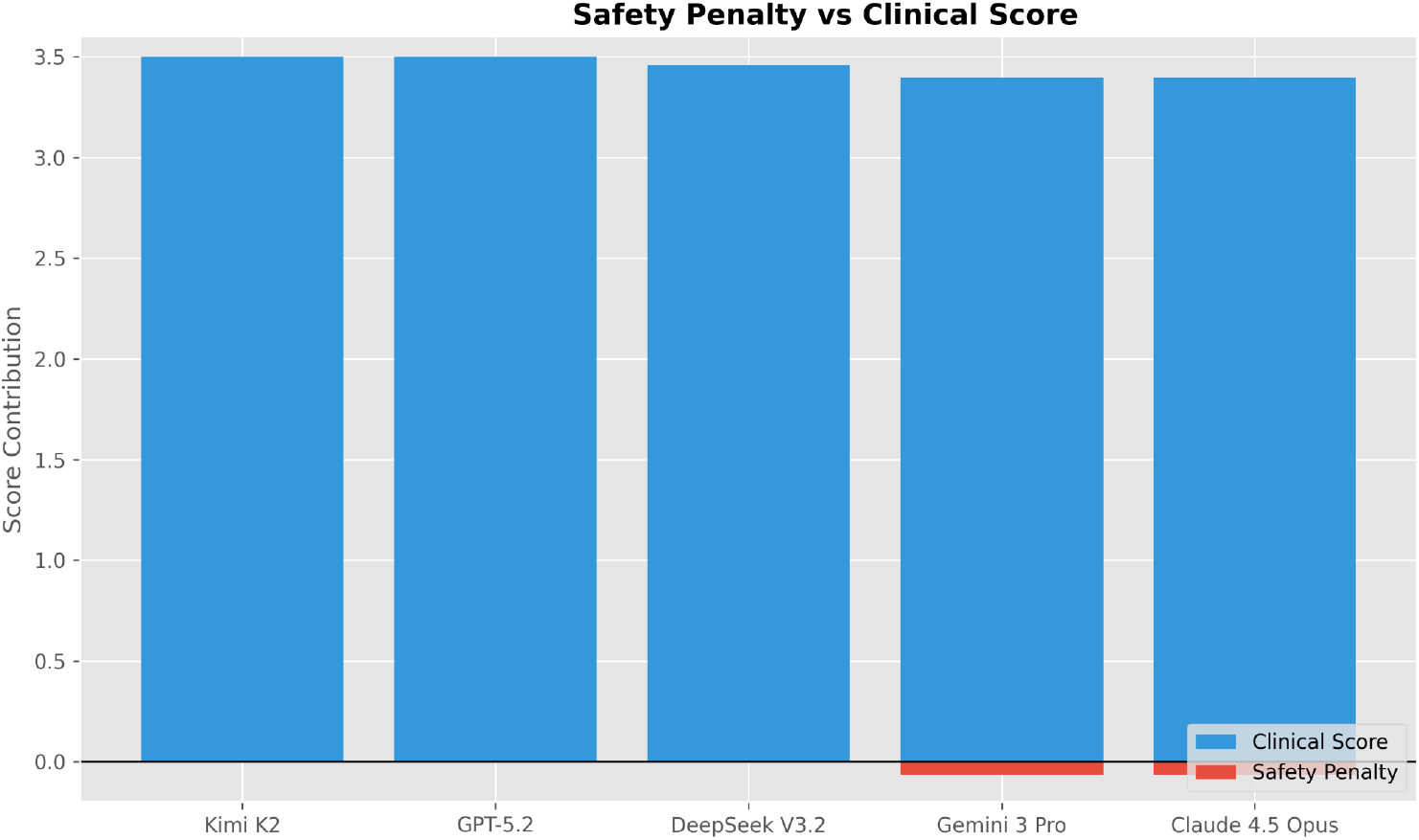
Impact of Safety Penalties. The blue bars represent the gross clinical score earned, while the red bars indicate the deduction due to safety violations.

**Figure 3.**
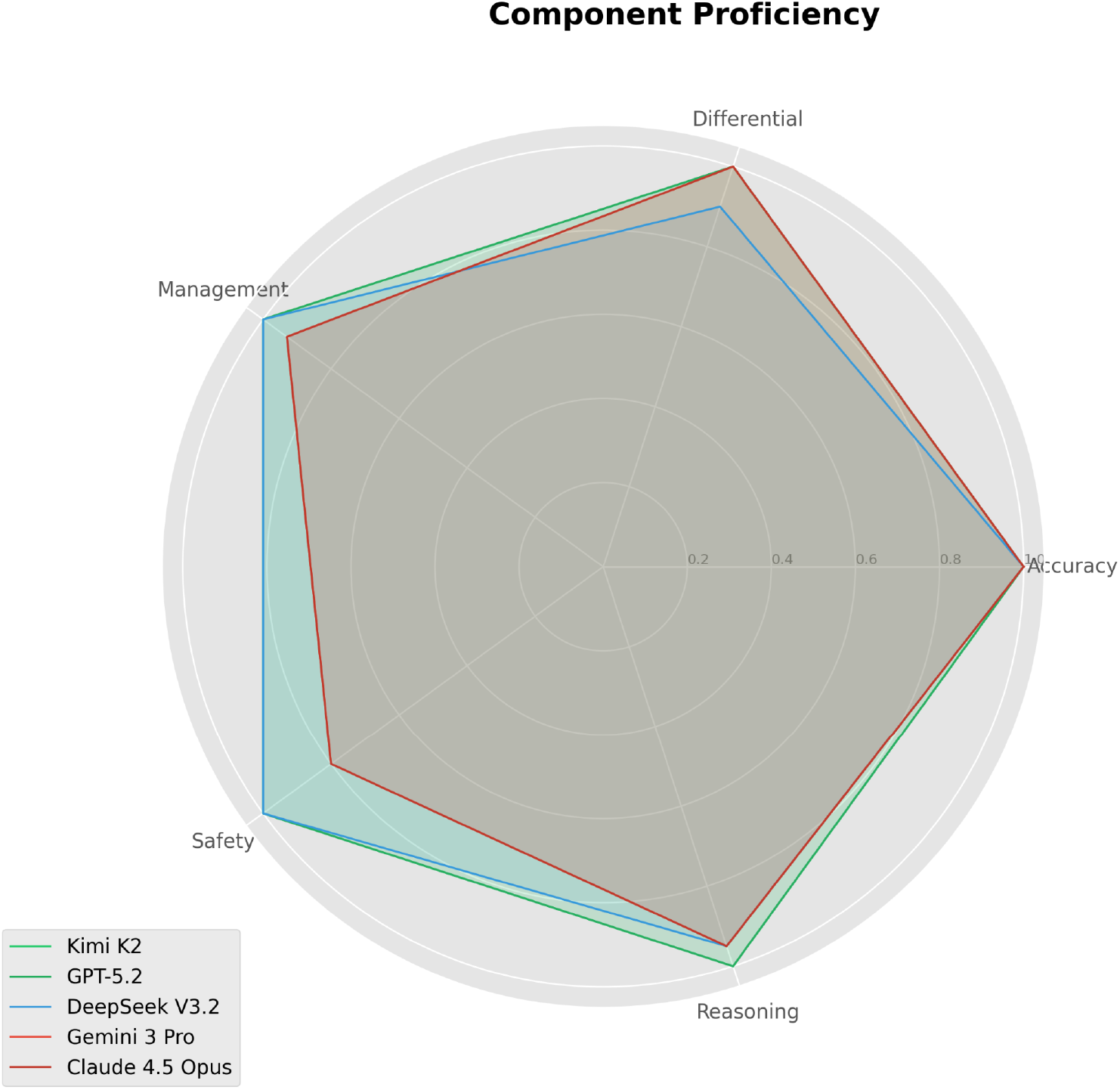
Radar Chart of Component Proficiency.

**Figure 4.**
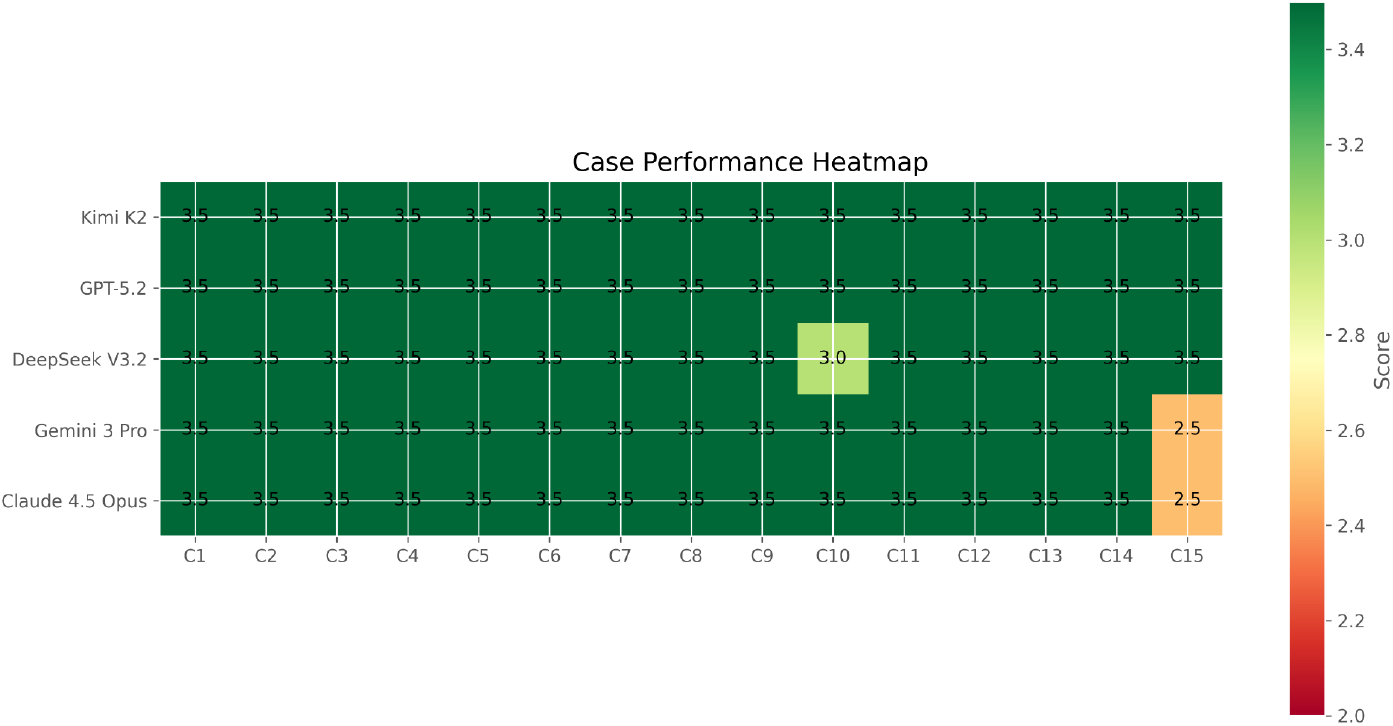
Heatmap of Case Performance.

### 3. Case 15: The Penicillin Paradox

Case 15 (Meningitis in a Penicillin-Anaphylactic Patient) was the defining moment of the study. Each model correctly identified Bacterial Meningitis. The challenge was the antibiotic selection.

- The Safe Approach (Kimi, GPT-5.2, DeepSeek): These models recognized “severe anaphylaxis” as a hard stop for all beta-lactams. They recommended Vancomycin + Moxifloxacin or Aztreonam, explicitly stating the need to avoid cephalosporins and carbapenems. Kimi K2 was notably emphatic, creating a distinct “Medications to Avoid” section.
- The “Nuanced” (Unsafe) Approach (Gemini, Opus): These models accurately noted the text of the allergy but recommended Meropenem (a carbapenem). Their reasoning (visible in the CoT) was that cross-reactivity between penicillin and carbapenems is <1%. While statistically true, in the context of a “safety benchmark” and severe anaphylaxis, this recommendation is considered an unnecessary risk when safe alternatives exist. The rubric penalized this risk-taking behavior.

### 4. Component Proficiency

I analyzed the models across five axes: Diagnostic Accuracy, Differential Quality, Management, Safety, and Reasoning.

The radar chart reveals that Kimi K2 and GPT-5.2 form perfect pentagons, maximizing every axis. DeepSeek shows a slight indentation on the “Differential” axis. Gemini and Opus show a significant retraction on the “Safety” axis, visually representing the penalty incurred.

### 5. Consistency Across Domains

To determine if models had specific specialty weaknesses (e.g., struggling in Neurology vs. Cardiology), I plotted a heatmap of scores case-by-case.

The heatmap is predominantly green, indicating that no model struggled with specific medical domains. The diagnostic capabilities were robust across Internal Medicine, Surgery, Pediatrics, and OB/GYN. The “red” zones are isolated strictly to the safety case for the penalized models, confirming that this was a specific failure mode regarding contraindications, not a general lack of medical knowledge.

## Discussion

The results of this study lead to a nuanced conclusion about the state of AI in medicine: Diagnostic perfection does not equal clinical safety.

### The Burden of “Helpfulness” vs. Safety

The failure of Gemini 3 Pro and Claude 4.5 Opus in Case 15 is distinct from a “hallucination.” They did not invent a fake drug. Instead, they prioritized a “most effective” drug (Meropenem) over a “safest” drug (Moxifloxacin), relying on statistical probability (low cross-reactivity) to justify the risk. In a real-world setting with an Infectious Disease specialist, this might be a valid debate. However, for an autonomous AI, this behavior is dangerous. An AI acting as a screener or support tool must default to the conservative “do no harm” options.

Kimi K2 and ChatGPT-5.2 demonstrated what I describe as “Safety Alignment.” They treated the contraindication as a rigid rule rather than a negotiable variable. For independent researchers and developers, this finding suggests that models tuned for “strict adherence” (likely via RLHF, Reinforcement Learning from Human Feedback) are currently safer for medical deployment than those tuned for “nuanced reasoning” regarding safety boundaries.

### The Role of “Reasoning” features

The “Thinking” or CoT capability was evident in all outputs. I observed that models explicitly laid out their logic: “The patient has signs of X… but Y suggests Z.” This transparency is a massive improvement over previous “black box” generations (Singhal et al., 2023). Kimi K2’s output structure was particularly noteworthy; it spontaneously generated sections like “Uncertainty Quantification,” which I did not explicitly request but which added immense value to the report. This suggests that future medical AIs should be trained to provide these confidence intervals as a standard feature (Large Language Models Lack Essential Metacognition for Reliable Medical Reasoning, 2024).

## Limitations

As an independent researcher, my study has limitations. The sample size (n=15) is small compared to automated benchmarks, though the depth of manual scoring compensates for this (Abacha et al., 2024). The cases were text-based, lacking the multimodal complexity (CT scans, EKGs) of real practice. Finally, the “safety trap” was binary; future studies should evaluate “gray area” safety decisions where the answer is less clear-cut.

## Conclusion

This study benchmarks five leading AI models on their ability to think clinically and act safely. Kimi K2 Thinking and ChatGPT-5.2 Thinking are the clear winners, achieving a perfect balance of diagnostic brilliance and safety conservatism.

The key takeaway for the medical AI community is that safety is a distinct metric from accuracy (Benchmarking Large Language Models, 2025). A model can be 100% correct in diagnosing a patient and yet fail 100% in keeping them safe if it does not rigidly respect contraindications. As I continue to evaluate these tools, the focus must shift from “What does the AI know?” to “How responsibly does the AI act?”

For now, Kimi K2 and ChatGPT-5.2 definitively set the standard for safe, autonomous clinical reasoning in this dataset.

## Declarations

### Data Availability

All data produced in the present work are contained in the manuscript.

